# Cost-effectiveness of paramedic administered ketamine compared to morphine for the management of acute severe pain from traumatic injury

**DOI:** 10.1101/2025.07.29.25332358

**Authors:** Kamran A Khan, Michael Smyth, Gavin D Perkins, Joyce Yeung, Alison Walker, Rebecca McLaren, Gordon Fuller, Stavros Petrou

## Abstract

**Background:** Pain after traumatic injury is common, yet few patients receive adequate pain relief. NHS paramedics have a limited formulary to treat severe pain.

**Objectives:** To estimate the cost-effectiveness of ketamine versus morphine for severe pain in acute traumatic injury.

**Methods:** A cost-utility analysis was conducted based on data from a pragmatic, multicentre, randomised controlled trial (PACKMAN). The base-case analysis took the form of an intention-to-treat analysis conducted from a UK National Health Service (NHS) and personal social services (PSS) perspective and separately from a societal perspective. Costs (£ 2021–2022 prices) were collected prospectively over a 6-month follow-up period. A bivariate regression of costs and quality-adjusted life-years (QALYs), with multiple imputation of missing data, was conducted to estimate the incremental cost per QALY gained and the incremental net monetary benefit (INMB) of ketamine in comparison to morphine. Sensitivity and pre-specified subgroup analyses explored uncertainty and heterogeneity in cost-effectiveness estimates.

**Results:** Participants (n=416) were randomised to ketamine (n=206) or morphine (n=210) amongst whom complete data for the economic evaluation was available for 189 (45.4%) participants. Mean (standard deviation [SD]) observed NHS and PSS costs over 6 months were £5,191 (£3,155) in the ketamine arm versus £5,143 (£3,897) in the morphine arm (mean difference [MD]: £47, p=0.926). Mean (SD) observed QALY estimates were 0.309 (0.10) versus 0.293 (0.010), respectively (MD: 0.016, p=0.273).

The base case (imputed) analysis generated an incremental cost of −£117 (95%CI: −£849 to £597) and incremental QALYs of 0.025 (95%CI: 0.010 to 0.041), indicating a 92%-96% probability of cost-effectiveness at cost-effectiveness thresholds of £20,000 and £30,000 per QALY. A sensitivity analysis, using observed data only (without imputation) generated an incremental cost of £233 (95%CI: −£783 to £1216) and incremental QALYs of 0.016 (95%CI: −0.013 to 0.044), indicating a lower 54%-62% probability of cost-effectiveness. The base-cost cost-effectiveness results remained robust to other sensitivity analyses.

**Conclusions:** This economic evaluation found that ketamine administered by paramedics to adults with severe pain following traumatic injuries is cost-effective compared to morphine. However, our results are subject to high levels of missing data, which were handled through recommended multiple imputation techniques.

## Background

Pain after traumatic injury is common, yet few patients receive adequate pain relief. At least 70% of ambulance calls involve patients experiencing pain.(1) NHS paramedics have a limited formulary to treat severe pain.(1) Observational studies suggest that current treatments leave many patients with inadequate pain relief in the prehospital environment.(2–6) The effective management of acute pain is important for humanitarian reasons, for improving patient experience and reducing adverse long term outcomes, such as chronic pain. In 2004, the World Health Organisation declared that effective management of pain is a universal human right. Poorly managed acute pain is also associated with increased chronic pain. Studies indicate chronic pain is common following trauma with a reported incidence of 15-30%, increasing to 62% in patients suffering major trauma.(7–9) Poorly managed postoperative pain leads to persistent pain in 10-50% of common surgeries, and that pain is severe in about 2-10% of these patients.(10) Military personnel injured in recent conflicts demonstrate a link between acute pain management and depression and post-traumatic stress disorder (PTSD). Early aggressive pain management exerts a protective effect on the development of PTSD (odd ratio (OR) 0.47 (95%CI 0.34-0.66) and depression (0.40 (95%CI 0.17 – 0.94).(11, 12) Provision of early and effective analgesia has the potential to reduce the risk of developing chronic pain and adverse mental health outcomes post trauma, which may in turn impact on patient’s long term quality of life.(13, 14)

A barrier to effective pain treatment is the limited formulary available to paramedics. The most frequently used drug for moderate to severe pain outside a hospital is morphine.(15) Yet morphine has several side effects (nausea, confusion, dizziness, drowsiness, respiratory depression, arrhythmia) that may limit its use.(16–19) This, and concerns about potential longer term dependence, limits effective use by clinicians.(20) Ketamine is perceived by many to be an ideal prehospital analgesic agent, favoured for its rapid onset of action, effective analgesia, good haemodynamic stability, and preservation of upper airway reflexes.(21) Ketamine has a distinct dose-response gradient in which smaller doses (0.1-0.3mg/kg) are analgesic and larger doses (2mg/kg) have an anaesthetic effect.(22) It exerts its effect by “disconnecting” the thalamocortical and limbic systems, effectively dissociating the central nervous system (CNS) from outside stimuli (e.g. pain, sight, sound).(23) Ketamine also stimulates the sympathetic nervous system and moderately increases heart rate and blood pressure. Ketamine does not affect respiration; patients breathe spontaneously and maintain airway control.(24) Furthermore, there is evidence to indicate that perioperative ketamine analgesia may prevent hyperalgesia, reducing the risk of developing persistent post-operative pain.(25, 26) This suggests the potential for ketamine analgesia to be associated with a lower incidence of chronic pain post trauma.

Due to its rapid onset and favourable side effect profile, ketamine is widely used in ambulance systems around the world.(27–32) In the UK, ketamine is currently restricted for use by prehospital doctors and a limited pool of specialist critical care paramedics (CCPs), targeted at the small number of cases needing critical care support.(33, 34) The lack of evidence and UK experience with ketamine limits access to a potentially effective treatment. Most trials of ketamine for analgesia have been small, of insufficient quality and were conducted in North America or Australia. (35–39) Patient expectations and approaches to health service delivery in these countries differ from the UK. No studies addressing the cost-effectiveness of ketamine for analgesia have been published. The National Institute for Health and Care Excellence (NICE) in the UK has identified the need for a pragmatic, randomised trial to determine the clinical and cost-effectiveness of ketamine against standard care (morphine)(40) This study therefore aimed to estimate the cost-effectiveness of ketamine for severe pain in acute traumatic injury when delivered by UK paramedics. The findings are intended to inform policy makers, guideline developers and ambulance services as to whether ketamine should be added to the paramedic formulary.

## Methods

### Trial background

The Paramedic Analgesia Comparing Ketamine and MorphiNe (PACKMAN) Trial was a pragmatic, multicentre, randomised, double blind randomised controlled trial (RCT) comparing the clinical and cost-effectiveness of ketamine versus morphine for severe pain in acute traumatic injury: the protocol has been published previously (41). In brief, acute trauma patients, aged 16 and over, who reported a pain score ≥7/10 on a 0-10 numeric rating scale (NRS) following acute traumatic injury, with Intravenous (IV) or intraosseous (IO) access, determined by a paramedic to require IV morphine or equivalent were eligible. The trial had a prespecified target sample size of 446 participants (41). Recruitment occurred between 10^th^ November 2021 and 16^th^ May 2023 from two large NHS ambulance services (West Midlands and Yorkshire NHS Ambulance Services) in England. The treatment intervention, ketamine, was supplied in ampoules containing 15mg in 1ml. The control intervention, morphine, was supplied in ampoules containing 10mg in 1 ml. The trial drugs were administered by slow IV (or IO) injection, titrated to effect over five minutes, aiming to give the minimal effective dose. If the patient continued to report pain 5 minutes after receiving the first full syringe (10ml), a second syringe was prepared and administered in a similar manner by the attending paramedic. A maximum of 20mls of trial drug could be administered, equating to a maximum dose of either 20mg morphine or 30mg ketamine. The ampoules were labelled as trial related investigational medicinal product (IMP) and paramedics were not able to identify which treatment they were administering.(41) Participants were randomised (1:1 ratio) to either ketamine or morphine. Numbered study drug packs in a pre-randomised sequence, were carried by participating ambulance paramedics. Randomisation occurred when the trial IMP pack was opened. The primary clinical outcome was the Sum of Pain Intensity Difference (SPID) assessed using a 0-10 numeric rating scale. Pain intensity was recorded prior to treatment administration and then at regular intervals following randomisation until arrival at hospital. Other important outcomes included overall pain relief, patient experience, tolerability, and the economic outcomes described below.

### Overview of economic analyses

The cost-utility analysis involved evaluation of economic costs, health-related quality of life (HRQoL) outcomes and cost-effectiveness of ketamine versus morphine where cost-effectiveness was expressed in terms of incremental cost per quality adjusted life year (QALY) gained. The base-case economic evaluation took the form of an intention-to-treat, imputed analysis conducted from a UK National Health Service (NHS) and personal social services (PSS) perspective in line with the NICE reference case.(42) The NHS payer perspective considers intervention-related treatment costs and other health service resource use and costs whilst a personal social services perspective includes services provided by local authorities for vulnerable groups, including older people. A 6-month time horizon for the economic evaluation was used mirroring the trial follow-up period and therefore no discounting was required.

### Costs

Three broad resource use and costs categories were delineated for cost estimation: (i) Direct intervention costs (medication costs); (ii) Direct healthcare and PSS (e.g. medications for side-effects, outpatient appointments, community health and social care) use during the 6 month follow-up; and (iii) for the purposes of a sensitivity analysis conducted from a societal perspective also included non-NHS & PSS costs (e.g. value of lost productivity, out of pocket expenses). All costs were expressed in pounds sterling and valued in 2021–22 prices. Where required, costs were inflated or deflated to 2021–22 prices using the NHS Cost Inflation Index (NHSCII).(43) The PACKMaN trial focused on administration of two alternative medications for pain relief in patients with severe pain. The intervention arm received ketamine hydrochloride whilst the control arm received morphine sulphate. The intervention components, how they were collected, associated resource use and source of unit costs are summarised in Supplementary Table 1 (Appendix). In according with NICE guidance, we captured NHS and PSS costs for both arms of the trial.(42) This included within-ambulance costs, inpatient care, outpatient care, community care, accident and emergency admission, medication, and personal social services. The methods for capturing the resource use and the sources for unit costs are outlined in Supplementary Table 2 (Appendix). Within ambulance costs were captured through the ambulance service data form, index admission costs were collected via the hospital data collection form, whilst the remaining health and social service resource use was collected through participant-completed questionnaires completed at 3 and 6 months post-randomisation. The identified resource inputs were valued using unit costs (Supplementary Table 3) identified through national cost compendia in accordance with NICE’s Guide to the Methods of Technology Appraisal.(42) Unit cost data were derived based on NHS England’s National schedule of NHS costs 2021-22 schedules, (44) the Personal social services research unit (PSSRU) Unit Costs of Health and Social Care 2022 compendium, (43) 2021-22 volumes of the British National Formulary,(45) NHS Supply Chain Catalogue 2021-22,(46) and the 2021-22 National Health Service Business Service Authority (NHSBSA) Prescription Cost Analysis (PCA) schedule.(47)Analyses from a societal perspective additionally encompassed economic values for work absences (by patients and their caregivers), travel costs and privately incurred health expenditures. These costs were self-reported by trial participants.

### Health⍰related quality of life outcomes

HRQoL were assessed using the EQ-5D-5L instrument, which defines HRQoL in terms of five dimensions (mobility, self-care, usual activities, pain/discomfort, anxiety/depression), each with five levels of severity.(48) For ethical, logistical and pragmatic reasons, it was not possible to capture baseline EQ-5D-5L measurements in patients suffering acute pain following trauma within this trial. This is not uncommon within trials involving emergency and critical care settings.(49) Ideally, the EQ-5D-5L would be completed at the time of randomisation or as soon as possible afterwards. This however was not possible in this trial. National age and gender specific norms for EQ-5D utility values were therefore applied at baseline.(50) HRQoL at 3 and 6 months post-randomisation was assessed using patient-completed EQ-5D-5L responses. Responses to the EQ-5D-5L descriptive system were mapped onto the EQ-5D-3L value set using the Alava HM et al. interim cross-walk algorithm,(51) as recommended by NICE in England and Wales. (42) Patient-level QALYs were estimated using the area under the curve approach, assuming linear interpolation between the utility scores, i.e., the preference-based values attached to the health states generated from the EQ-5D-5L descriptive system.

### Handling of missing data

Multiple imputation by chained equations was used to predict missing costs and health utility scores based on the assumption that data were missing at random (MAR). The MAR assumption was tested through a series of logistic regression analyses comparing participants’ characteristics for those with and without missing endpoint data. Imputation was achieved using predictive mean matching, which has the advantage of preserving nonlinear relationships and correlations between variables within the data. Fifty imputed datasets were generated to inform the base-case and subsequent sensitivity and subgroup analyses. Parameter estimates were pooled across the imputed datasets using Rubin’s rules to account for between- and within-imputation components of variance terms associated with parameter estimates.(52)

### Cost⍰effectiveness analysis

Mean resource use, cost and health utility values were compared between the trial arms using two sample t-tests. Mean incremental costs and mean incremental QALYs were estimated using seemingly unrelated regression (SUR) methods that account for the correlation between costs and outcomes.(53) Differences between groups, along with confidence intervals (CIs), were estimated using non-parametric bootstrap estimates (10,000 replications) of regression models. The cost equation was adjusted using: type of ambulance service (West Midland Ambulance Service (WMAS), Yorkshire Ambulance Service (YAS)), age category (<60, ≥60), gender (male, female), administration of IV analgesia prior to randomisation (Yes, No), and weight ((i) >0 and <70, ii) ≥70 and <85, iii) ≥85 kgs). The QALY equation was adjusted using baseline utilities, ambulance service (WMAS, YAS), age category (<60, ≥60), gender (male, female), administration of IV analgesia prior to randomisation (Yes, No), and weight ((i) >0 and <70, ii) ≥70 and <85, iii) ≥85 kgs)). Following imputation, bootstrapping was used to generate the joint distribution of costs and outcomes and to populate a cost-effectiveness plane. The incremental cost-effectiveness ratio (ICER) for ketamine was estimated by dividing the between-group difference in adjusted mean total costs by the between-group difference in adjusted mean QALYs. Mean ICER values were compared against cost-effectiveness threshold values (i.e. society’s willingness to pay for an additional QALY) ranging between £20,000 and £30,000 per QALY gained in line with NICE guidance. (42) ICER values lower than the threshold are considered cost-effective for use in the UK NHS. The incremental net monetary benefit (INMB) of switching from morphine to ketamine was also calculated at each of these cost-effectiveness threshold values. The net monetary benefit is the economic benefit of an intervention (expressed in monetary terms) net of all costs. A positive incremental NMB suggests that, on average, ketamine is cost-effective compared with morphine, at the given cost-effectiveness threshold.

### Sensitivity and subgroup analyses

Pre-specified sensitivity analyses were undertaken to assess the impact of uncertainty surrounding components of the economic evaluation and included restricting the analyses to complete cases (i.e. the sample of participants with no missing costs or outcome data at any time point), replicating the analysis from a societal perspective, and changing the baseline utility assumption (assumed a fixed utility of 0 for everyone). Prespecified subgroup analyses were conducted by age category (<60, ≥60), gender (male, female), administration of IV analgesia prior to randomisation ((Yes, No), weight (i) >0 and <70, ii) ≥70 and <85, iii) ≥85 kgs). In addition, a scenario analysis was conducted estimating the incremental cost per score point reduction in the sum of pain intensity difference (SPID) the time horizon for this was constrained to the period between randomisation and initial hospital discharge.

## Results

### Study population and data completeness

Baseline characteristics of participants were well-matched between the randomised groups (Table 1). Complete QALY profiles were available for 196 (47%) participants based on the EQ-5D-5L (Table 2). Completion of resource use data for the economic evaluation was similar (53%-57%) at each time-point between the ketamine and morphine groups (Table 2). There were no significant differences in the sociodemographic characteristics between participants with or without complete data (Supplementary Table 4).

**Table 1:** Baseline characteristics by trial arm.

|  | Ketamine (n=206) | Morphine (n=210) |
| --- | --- | --- |
| Baseline characteristics |  |  |
| Ambulance service (n, %) |  |  |
| WMAS | 107 (51.9%) | 109 (51.9%) |
| YAS | 99 (48.1%) | 101 (48.1%) |
| Age category (n, %) |  |  |
| <60 | 85 (41.3%) | 82 (39.0%) |
| ≥60 | 121 (58.7%) | 128 (61.0%) |
| Gender (n, %) |  |  |
| Female | 110 (53.4%) | 110 (52.4%) |
| Male | 96 (46.6%) | 100 (47.6%) |
| Analgesia<sup>1</sup>(n, %) |  |  |
| No | 119 (57.8%) | 122 (58.1%) |
| Yes | 87 (42.2%) | 88 (41.9%) |
| Weight category (n, %) |  |  |
| >0 and <70 | 72 (35.0%) | 62 (29.5%) |
| ≥70 and <85 | 71 (34.5%) | 69 (32.9%) |
| ≥85 | 63 (30.5%) | 79 (37.6%) |
| Baseline utilities<sup>2</sup> (mean (SD)) | 0.7809 (0.07) | 0.7818 (0.08) |
| Baseline Pain Score | 8.8358 (1.19) | 8.8469 (1.21) |
| <sup>1</sup>Administration of IV analgesia prior to randomisation. |  |  |
| <sup>2</sup>Age and gender specific population norm values. |  |  |

**Table 2:**
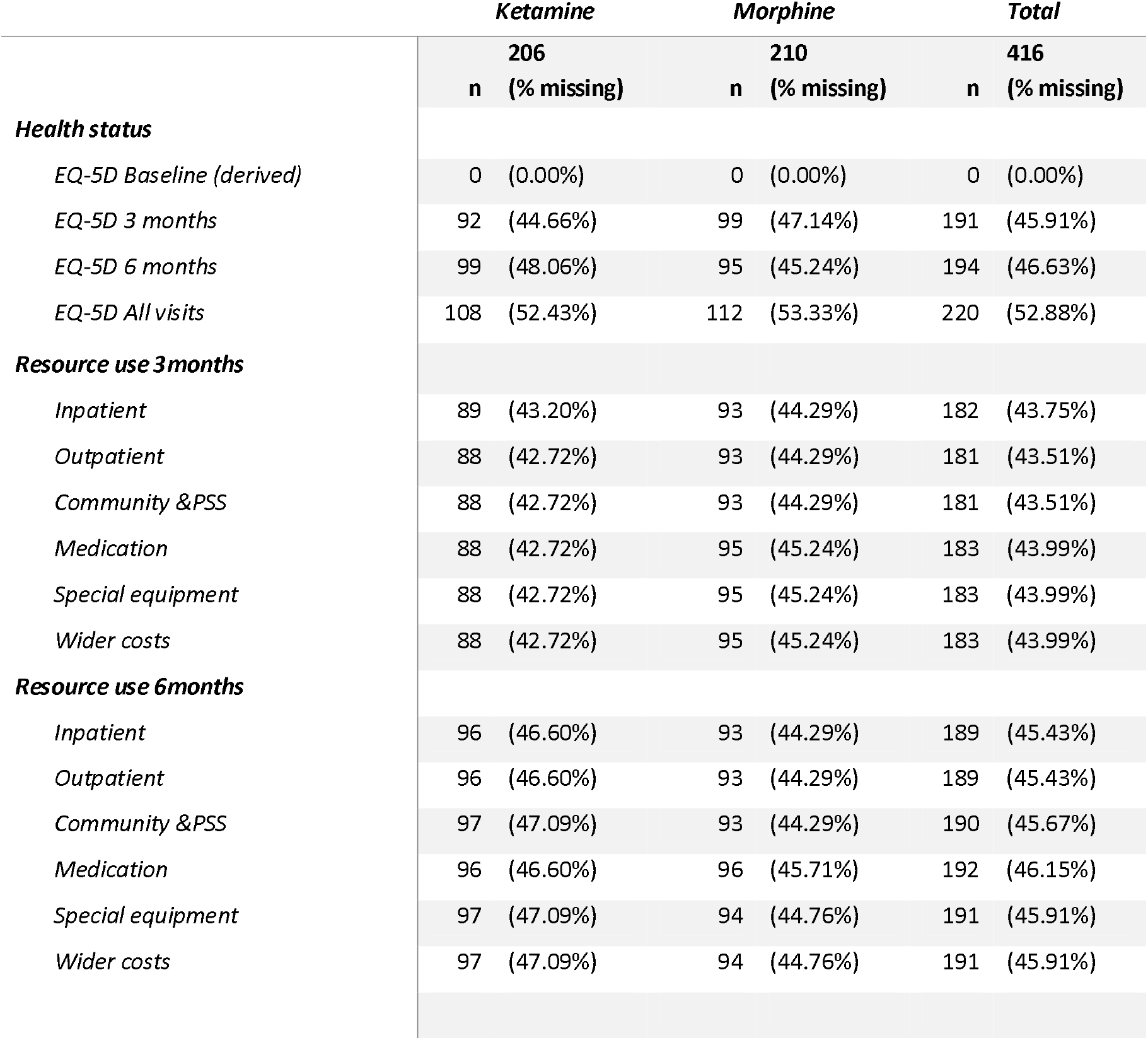
Missingness of data by follow-up visit.

|  | *Ketamine* |  | *Morphine* |  | *Total* |  |
| --- | --- | --- | --- | --- | --- | --- |
|  | **206** |  | **210** |  | **416** |  |
|  | **n** | **(% missing)** | **n** | **(% missing)** | **n** | **(% missing)** |
| ***Health status*** |  |  |  |  |  |  |
| *EQ-5D Baseline (derived)* | 0 | (0.00%) | 0 | (0.00%) | 0 | (0.00%) |
| *EQ-5D 3 months* | 92 | (44.66%) | 99 | (47.14%) | 191 | (45.91%) |
| *EQ-5D 6 months* | 99 | (48.06%) | 95 | (45.24%) | 194 | (46.63%) |
| *EQ-5D All visits* | 108 | (52.43%) | 112 | (53.33%) | 220 | (52.88%) |
| ***Resource use 3months*** |  |  |  |  |  |  |
| *Inpatient* | 89 | (43.20%) | 93 | (44.29%) | 182 | (43.75%) |
| *Outpatient* | 88 | (42.72%) | 93 | (44.29%) | 181 | (43.51%) |
| *Community & PSS* | 88 | (42.72%) | 93 | (44.29%) | 181 | (43.51%) |
| *Medication* | 88 | (42.72%) | 95 | (45.24%) | 183 | (43.99%) |
| *Special equipment* | 88 | (42.72%) | 95 | (45.24%) | 183 | (43.99%) |
| *Wider costs* | 88 | (42.72%) | 95 | (45.24%) | 183 | (43.99%) |
| ***Resource use 6months*** |  |  |  |  |  |  |
| *Inpatient* | 96 | (46.60%) | 93 | (44.29%) | 189 | (45.43%) |
| *Outpatient* | 96 | (46.60%) | 93 | (44.29%) | 189 | (45.43%) |
| *Community & PSS* | 97 | (47.09%) | 93 | (44.29%) | 190 | (45.67%) |
| *Medication* | 96 | (46.60%) | 96 | (45.71%) | 192 | (46.15%) |
| *Special equipment* | 97 | (47.09%) | 94 | (44.76%) | 191 | (45.91%) |
| *Wider costs* | 97 | (47.09%) | 94 | (44.76%) | 191 | (45.91%) |

### Cost of intervention

Mean total intervention costs are presented for each group (Supplementary Table 5). These varied between £21.76 (ketamine) and £23.89 (morphine). The information on cost components can be found in Supplementary Table 3.

### Resource utilisation

For health and personal social service use, shown in Supplementary Table 5, there were non-significant differences between the two groups in utilisation of hospital inpatient and outpatient care. In terms of community-based health and social care, there were significantly higher visits to the GP for the ketamine arm (mean (SD) 2.45 (1.79)) vs the morphine arm (mean (SD) 1.50 (0.79)). For all other categories of community-based health and social care, there were non-significant differences between the two groups in resource utilisation.

### Total economic costs

For the base-case (imputed) analysis, mean NHS and PSS costs, inclusive of intervention costs, over the entire follow-up period were £5207 for the ketamine arm versus £5324 for the morphine arm (Supplementary Table 6). There was a non-significant incremental cost saving in the ketamine arm of £117. Mean total societal costs, for the entire follow-up period, inclusive of the intervention cost, were £6266 in the ketamine arm compared with £6373 in the morphine group (Supplementary Table 6). This generated a non-significant incremental cost increase of £107 in favour of the ketamine arm. The estimates of economic costs for non-imputed (complete) cases are shown in Supplementary Table 5 and follow the same pattern as the imputed base case analysis.

### Health-related quality of life outcomes

For the base-case analysis, mean (SE) participant reported QALY estimates for the entire period were 0.314 (0.01) for the ketamine arm versus 0.289 (0.01) for the morphine arm; the mean between group difference was 0.0253 (Supplementary Table 6).

### Cost-effectiveness results: base-case analysis (imputed costs and adjusted

The base-case economic evaluation (NHS and PSS perspective, imputed costs and QALYs and adjusted for covariates) indicated that ketamine was associated with non-significant lower NHS and PSS costs (−£117, 95% CI − £849 to £597) and a significant improvement in QALYs (0.025, 95% CI 0.010 to 0.041). The mean ICER for ketamine was estimated at −£4,610 (southeast quadrant) per QALY gained, i.e. on average, ketamine was associated with a lower cost and an improvement in health. The associated mean INMB at cost-effectiveness thresholds of £20,000 and £30,000 per QALY were £631 and £884, respectively (Table 3). The base-case mean INMB was>0, suggesting that the use of ketamine would result in an average net economic gain. The probability of cost-effectiveness for ketamine was estimated as 92% and 96% at cost-effectiveness thresholds of £20,000 and £30,000 per QALY, respectively. The joint distribution of costs and outcomes for the base-case analysis is presented graphically in Fig. 1. The cost-effectiveness acceptability curve is shown in Fig. 2.

**Table 3:**
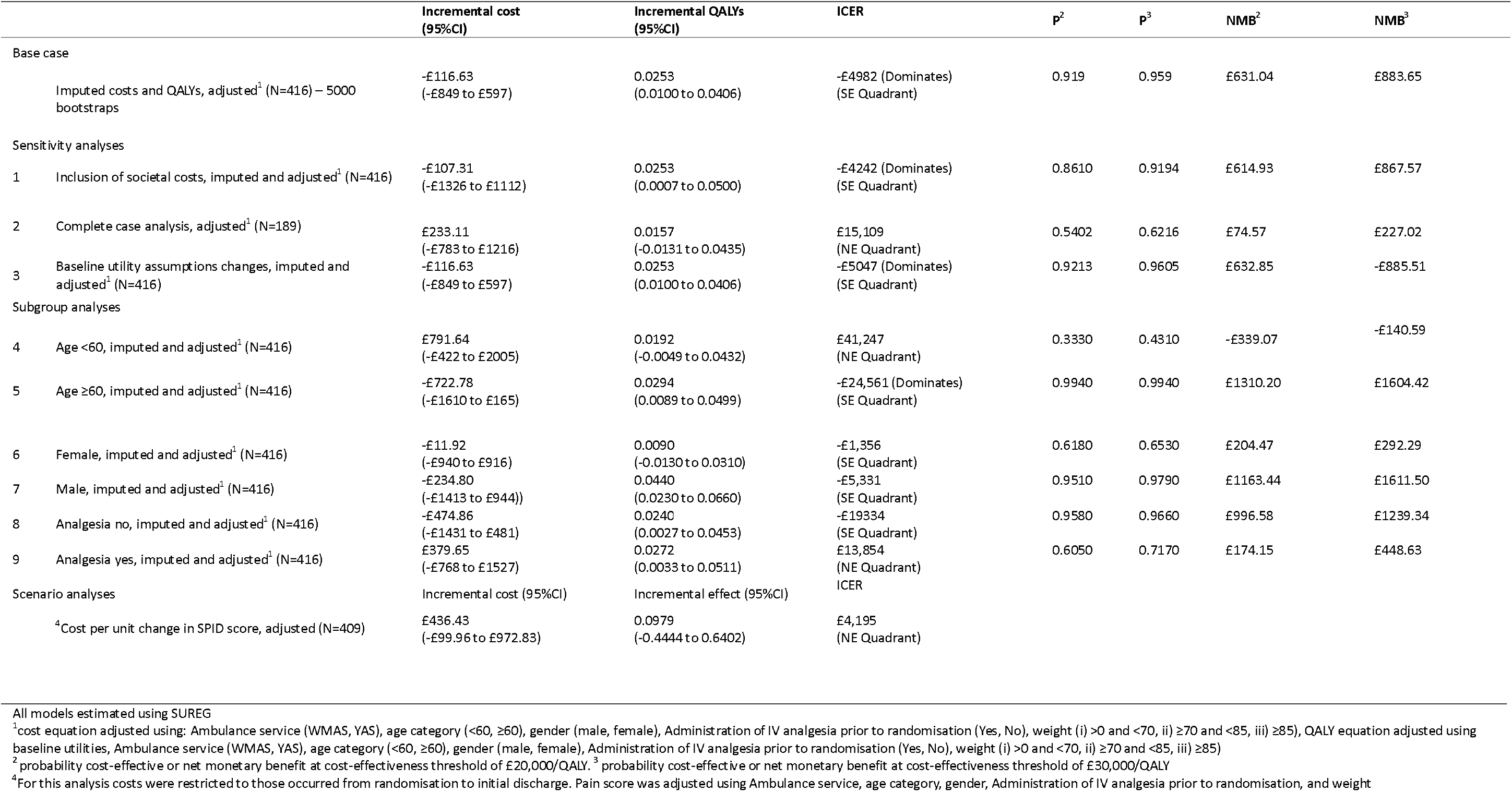
Cost-effectiveness results.

|  |  | Incremental cost<br>(95%CI) | Incremental QALYs<br>(95%CI) | ICER | p<sup>2</sup> | p<sup>3</sup> | NMB<sup>2</sup> | NMB<sup>3</sup> |
| --- | --- | --- | --- | --- | --- | --- | --- | --- |
| Base case |  |  |  |  |  |  |  |  |
|  | Imputed costs and QALYs, adjusted<sup>1</sup> (N=416) – 5000 bootstraps | -£116.63<br>(-£849 to £597) | 0.0253<br>(0.0100 to 0.0406) | -£4982 (Dominates)<br>(SE Quadrant) | 0.919 | 0.959 | £631.04 | £883.65 |
| Sensitivity analyses |  |  |  |  |  |  |  |  |
| 1 | Inclusion of societal costs, imputed and adjusted<sup>1</sup> (N=416) | -£107.31<br>(-£1326 to £1112) | 0.0253<br>(0.0007 to 0.0500) | -£4242 (Dominates)<br>(SE Quadrant) | 0.8610 | 0.9194 | £614.93 | £867.57 |
| 2 | Complete case analysis, adjusted<sup>1</sup> (N=189) | £233.11<br>(-£783 to £1216) | 0.0157<br>(-0.0131 to 0.0435) | £15,109<br>(NE Quadrant) | 0.5402 | 0.6216 | £74.57 | £227.02 |
| 3 | Baseline utility assumptions changes, imputed and adjusted<sup>1</sup> (N=416) | -£116.63<br>(-£849 to £597) | 0.0253<br>(0.0100 to 0.0406) | -£5047 (Dominates)<br>(SE Quadrant) | 0.9213 | 0.9605 | £632.85 | -£885.51 |
| Subgroup analyses |  |  |  |  |  |  |  |  |
| 4 | Age <60, imputed and adjusted<sup>1</sup> (N=416) | £791.64<br>(-£422 to £2005) | 0.0192<br>(-0.0049 to 0.0432) | £41,247<br>(NE Quadrant) | 0.3330 | 0.4310 | -£339.07 | -£140.59 |
| 5 | Age ≥60, imputed and adjusted<sup>1</sup> (N=416) | -£722.78<br>(-£1610 to £165) | 0.0294<br>(0.0089 to 0.0499) | -£24,561 (Dominates)<br>(SE Quadrant) | 0.9940 | 0.9940 | £1310.20 | £1604.42 |
| 6 | Female, imputed and adjusted<sup>1</sup> (N=416) | -£11.92<br>(-£940 to £916) | 0.0090<br>(-0.0130 to 0.0310) | -£1,356<br>(SE Quadrant) | 0.6180 | 0.6530 | £204.47 | £292.29 |
| 7 | Male, imputed and adjusted<sup>1</sup> (N=416) | -£234.80<br>(-£1413 to £944)) | 0.0440<br>(0.0230 to 0.0660) | -£5,331<br>(SE Quadrant) | 0.9510 | 0.9790 | £1163.44 | £1611.50 |
| 8 | Analgesia no, imputed and adjusted<sup>1</sup> (N=416) | -£474.86<br>(-£1431 to £481) | 0.0240<br>(0.0027 to 0.0453) | -£19334<br>(SE Quadrant) | 0.9580 | 0.9660 | £996.58 | £1239.34 |
| 9 | Analgesia yes, imputed and adjusted<sup>1</sup> (N=416) | £379.65<br>(-£768 to £1527) | 0.0272<br>(0.0033 to 0.0511) | £13,854<br>(NE Quadrant) | 0.6050 | 0.7170 | £174.15 | £448.63 |
| Scenario analyses |  | Incremental cost (95%CI) | Incremental effect (95%CI) | ICER |  |  |  |  |
|  | <sup>4</sup>Cost per unit change in SPID score, adjusted (N=409) | £436.43<br>(-£99.96 to £972.83) | 0.0979<br>(-0.4444 to 0.6402) | £4,195<br>(NE Quadrant) |  |  |  |  |

All models estimated using SUREG
<sup>1</sup>cost equation adjusted using: Ambulance service (WMAS, YAS), age category (<60, ≥60), gender (male, female), Administration of IV analgesia prior to randomisation (Yes, No), weight (i) >0 and <70, ii) ≥70 and <85, iii) ≥85), QALY equation adjusted using baseline utilities, Ambulance service (WMAS, YAS), age category (<60, ≥60), gender (male, female), Administration of IV analgesia prior to randomisation (Yes, No), weight (i) >0 and <70, ii) ≥70 and <85, iii) ≥85)
<sup>2</sup>probability cost-effective or net monetary benefit at cost-effectiveness threshold of £20,000/QALY. <sup>3</sup> probability cost-effective or net monetary benefit at cost-effectiveness threshold of £30,000/QALY
<sup>4</sup>For this analysis costs were restricted to those occurred from randomisation to initial discharge. Pain score was adjusted using Ambulance service, age category, gender, Administration of IV analgesia prior to randomisation, and weight

**Figure 1:**
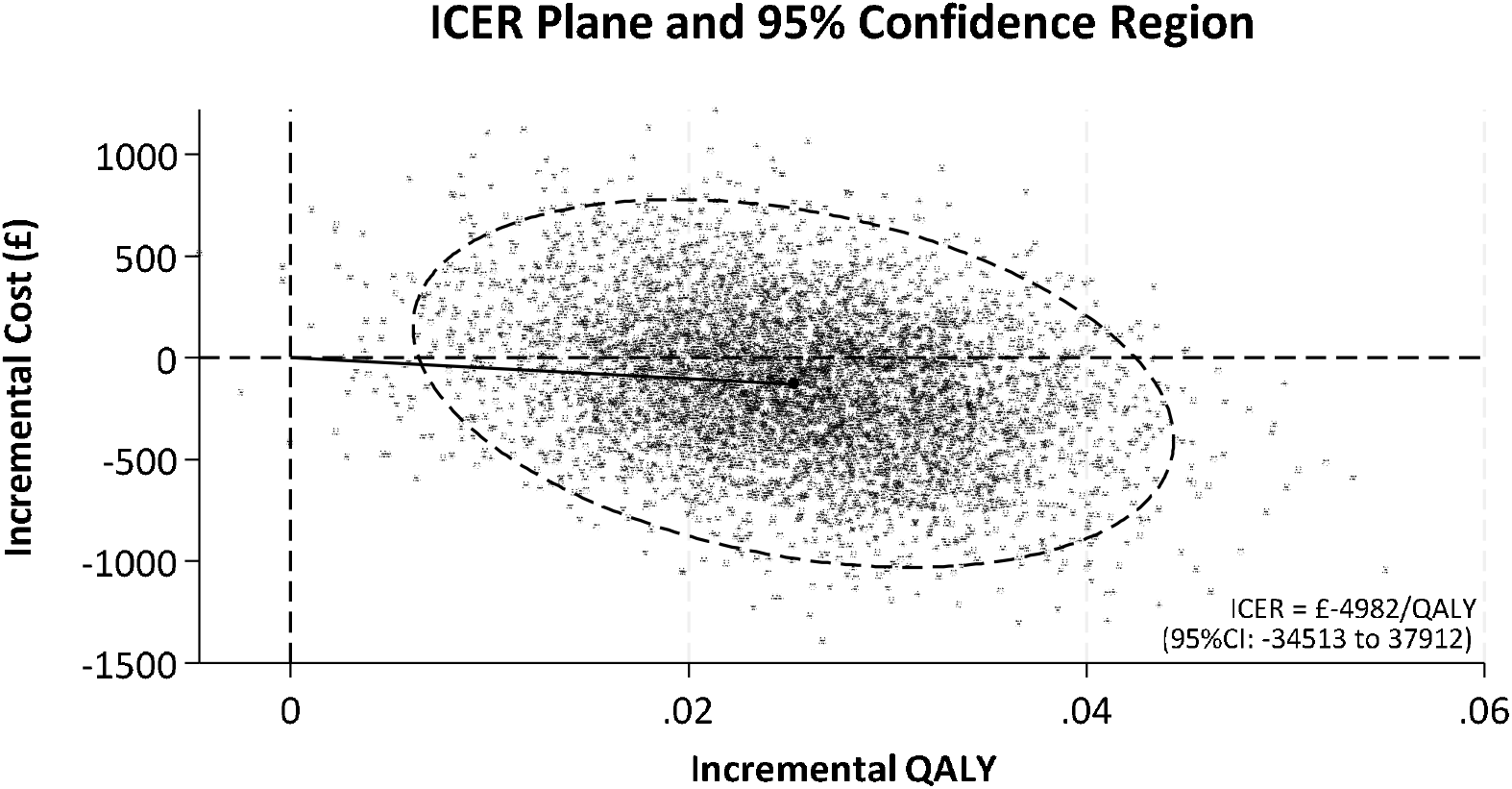
Cost-effectiveness plane, base case (Imputed costs and QALYs, adjusted)

**Figure 2:**
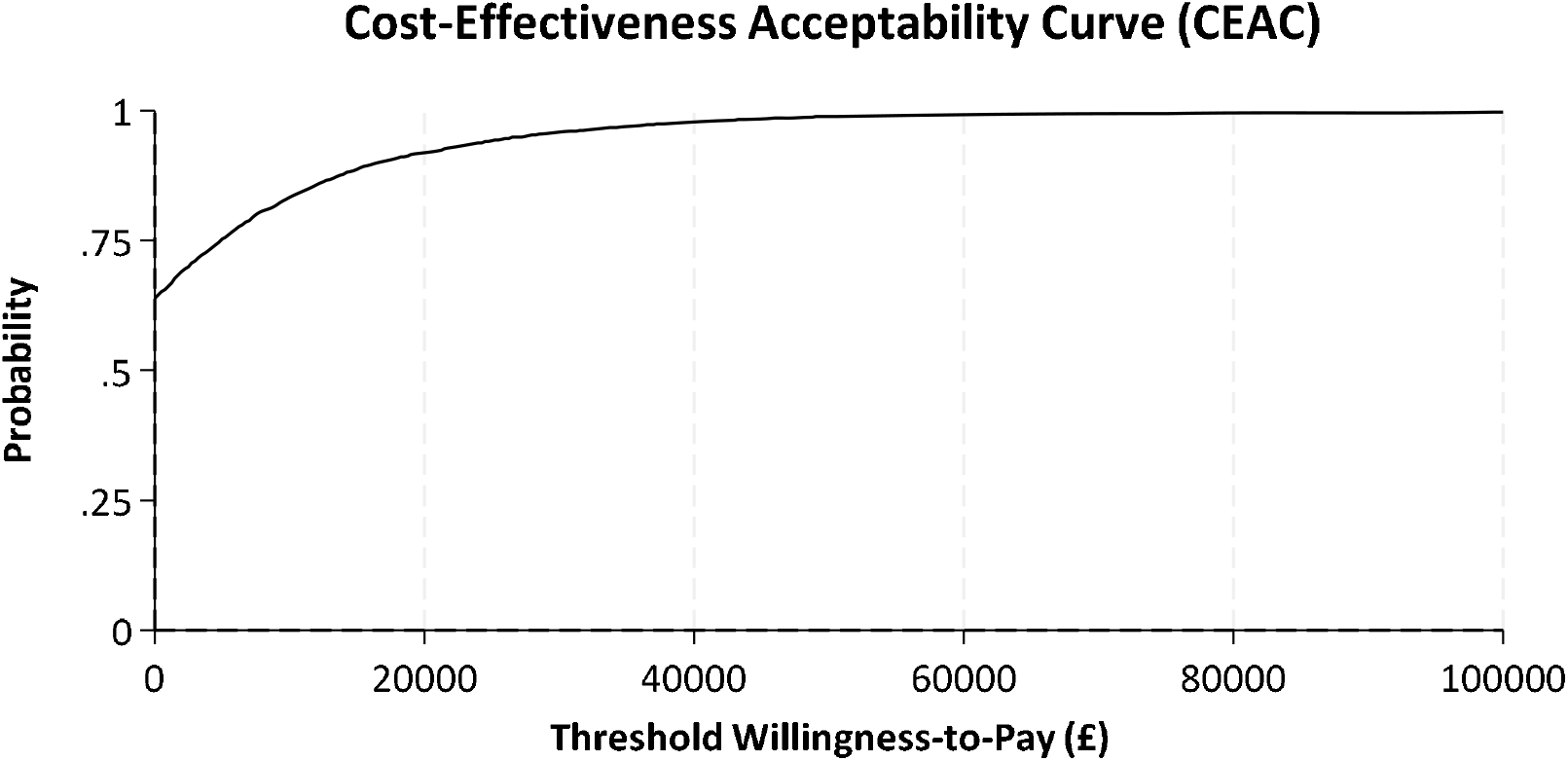
Cost-Effectiveness Acceptability Curve (CEAC), base case (Imputed costs and QALYs, adjusted)

### Sensitivity and subgroup analyses

The sensitivity analysis conducted from a societal perspective found a similar probability that ketamine was cost-effective of between 86 and 92% across cost-effectiveness thresholds (Table 3). The sensitivity analysis based on complete cases showed that the cost and QALY difference was non-significant and the probability that ketamine was cost-effective decreased to between 54 and 62% across cost-effectiveness thresholds. Using a baseline utility of 0 for all participants did not impact the results.

The pre-planned subgroup analyses suggested that ketamine was more cost-effective in the following subgroups: participants aged ≥60, males, and participants that did not receive IV analgesia prior to randomisation (Table 3), although the interaction terms in the underlying regression models were not significant.

The scenario analysis estimating the cost per unit change in SPID score indicated that ketamine was associated with non-significant increase in costs from randomisation to initial discharge from hospital (£436, 95% CI − £100 to £973) and a non-significant reduction in total pain (0.0979, 95% CI −0.444 to 0.640). The mean ICER for ketamine was estimated at £4,195 (northeast quadrant) per unit pain score reduction, i.e. on average, ketamine was associated with a higher cost and a reduction in pain score.

## Discussion

This trial-based economic evaluation revealed that the use of ketamine led, on average, to a modest increase in health-related quality of life, without increased cost, over a 6-month follow-up period. The resulting ICER from an NHSS and PSS perspective falls favourably below the recommended NICE cost-effectiveness threshold of £20,000 per QALY though the uncertainty around the mean ICER was large. From a societal perspective, ketamine was similarly cost-effective. There was no difference in clinical effectiveness (pain relief) when compared to morphine from randomisation to arrival at hospital.

There were some challenges when analysing the trial data, including persistent missingness at both follow up points, an imbalance of missingness by ambulance service, and a bimodal pattern of costs in both treatment arms. To address the missing data, imputation was carried out in accordance with the pre-specified health economic analysis plan. The robustness of the imputation was explored by varying the imputation seed and number of (discarded) burn-ins: the results were stable. Burn in traces were checked for adequate mixing and adequacy of the Markov chain Monte Carlo (MCMC) process. The number of draws used for the imputation was 50, this was adequate when checked against the uppermost fraction of missing information (FMI), which was 40%. There is no formal way of checking if the data are missing not-at-random (MNAR), but variables were identified that predicted missingness and included in the imputation model. A seemingly unrelated regression model was used for the base case analysis as it features the natural scale of the data and assumes normality of the bootstrap estimates for sample means. The distribution family for the dependent variables was explored and a gamma distribution with log link was found to improve the cost model specification, while the gaussian distribution was retained for the QALY variable. To preserve a bivariate analysis, a version of the base case was run using generalized structural equation modelling (GSEM) producing statistically similar findings. Several covariates in the base case model were significant. These were explored to see if they interacted with treatment where a significant interaction would suggest varying cost-effectiveness for the interaction sub-groups.

Our imputed analyses of cost-effectiveness outcomes gave a more optimistic estimate, reflecting some adjustment for the patterns of missingness. The evidence of HRQoL benefits adds to the emerging evidence base from clinical trials that demonstrate improvements in pain from ketamine. (35–39) Without economic modelling beyond the current parameters of the trial, the longer-term cost-effectiveness of ketamine cannot be ascertained.

We noted that the intervention appears less cost effective in participants who were younger, required analgesia prior to randomisation and in females. As with all sub-group analyses, these should be considered exploratory only, and our primary estimates account for all people. We used a pragmatic approach to sampling, and hence our findings should be generalisable. To the best of our knowledge there is no comparable evidence for cost-effectiveness of ketamine in trauma patients in the broader literature.

Strengths of the current economic evaluation are that the trial was prospectively designed for a cost-effectiveness analysis using individual-level data to reach a confirmatory conclusion. There are some limitations to this economic evaluation. Firstly, utility measurements were collected at only two time-points (3 months and 6 months) post-randomisation. Evidence suggests that the timing of assessment can significantly influence cost-effectiveness results when using the EQ-5D, particularly when participants experience recurrent health fluctuations. (54) In such cases, the linear interpolation of utility data may fail to reflect HRQoL fluctuations over short periods and the uncertainty is compounded by missing data. While the trial may have captured differences in chronic pain, it may have missed changes in acute pain occurring before the three-month follow-up. Secondly, resource use data were retrospectively recalled by participants, and this could have led to recall bias, though we cannot predict the direction of this bias. Findings form literature are mixed, suggesting that resource use may be under-reported, over-reported or they may be good agreement between patient/carer recall and data extracted from medical records, depending on how well the resource use measures are structured.(55) Because the recall periods and questionnaires were standardised across randomised groups, retrospective recall is unlikely to have biased results in favour of one group. Thirdly, our approaches to collecting resource use data did not disentangle resource use associated with trauma from resource use associated with broader health factors. Fourthly, there were high levels of missingness in the study data. However, we handled missingness within the health economic data through recommended multiple imputation techniques that address the inherent biases associated with estimating effects on the basis of complete data.

## Conclusions

In this economic evaluation based upon a randomised controlled trial, ketamine administered by paramedics to adults with severe pain following traumatic injuries was cost-effective compared to morphine.

## Supporting information

CHEERS 2022 Checklist

## List of abbreviations

CCPs: Critical care paramedics
CI: Confidence intervals
CNS: Central nervous system
FMI: Fraction of missing information
GSEM: Generalized structural equation modelling
HRQoL: Health-related quality of life
ICER: Incremental cost-effectiveness ratio
IMP: Investigational medicinal product
INMB: Incremental net monetary benefit
IO: Intraosseous
IV: Intravenous
MAR: Missing at random
MCMC: Markov chain Monte Carlo
MD: Mean difference
MNAR: Missing not-at-random
NHS: National health service
NHSBSA: National Health Service Business Service Authority
NHSCII: NHS Cost Inflation Index
NICE: National Institute for Health and Care Excellence
NRS: Numeric rating scale
OR: Odd ratio
PACKMAN: Paramedic Analgesia Comparing Ketamine and MorphiNe
PCA: Prescription Cost Analysis
PSS: Personal social services
PSSRU: Personal social services research unit
PTSD: Post-traumatic stress disorder
QALY: Quality-adjusted life-year
RCT: Randomised controlled trial
SD: Standard deviation
SPID: Sum of Pain Intensity Difference
SUR: Seemingly unrelated regression
WMAS: West midland ambulance service
YAS: Yorkshire ambulance service

## Declarations

### Ethics approval and consent to participate

Ethics approval for the PACKMAN trial was given by West of Scotland Research Ethics Committee (REC number 16/LO/0349) on 01/09/2020.

### Consent for publication

Not applicable.

### Availability of data and materials

The datasets analysed during the current study are available from the corresponding author upon reasonable request.

### Author contributions

CRediT author statement

**Kamran Khan:** Methodology, Formal analysis, Writing – Original draft

**Michael Smyth**: Conceptualization, Funding acquisition, Writing - Review & Editing

**Gavin Perkins**: Conceptualization, Funding acquisition, Writing - Review & Editing

**Joyce Yeung**: Conceptualization, Funding acquisition, Writing - Review & Editing

**Alison Walker**: Conceptualization, Funding acquisition, Writing - Review & Editing

**Rebecca McLaren**: Conceptualization, Funding acquisition, Writing - Review & Editing

**Gordon Fuller**: Conceptualization, Funding acquisition, Writing - Review & Editing

**Stavros Petrou**: Conceptualization, Funding acquisition Writing - Review & Editing, Supervision

## Acknowledgements

SP receives support as a National Institute for Health and Care Research (NIHR) Senior Investigator (NF-SI-0616-10103) and from the NIHR Applied Research Collaboration Oxford and Thames Valley.

GDP is supported by the National Institute for Health Research (NIHR) Applied Research Collaboration (ARC) West Midlands.

The views expressed are those of the author(s) and not necessarily those of the NIHR or the Department of Health and Social Care

