## Appendix for "Cost-effectiveness of paramedic administered ketamine compared to morphine for the management of acute severe pain from traumatic injury"

### Supplementary Information

**Table S1: Direct intervention resource use and cost sources**

| ***Intervention arm*** | | | |
| --- | --- | --- | --- |
| **Resource type** | **Resource use** | **How collected** | **Unit costs source** |
| Ketamine hydrochloride | Number and ml of doses administered. | Recorded within ambulance service data | BNF 2021/22(45)  NHSSC 2021/22(46) |
| ***Control arm*** | | | |
| Morphine sulphate | Number and ml of doses administered. | Recorded within ambulance service data | BNF 2021/22(45)  NHSSC 2021/22(46) |

**Table S2: Sources of health and social care costs for both trial arms**

| **Resource type** | **Resource use** | **How collected** | **Unit cost sources** |
| --- | --- | --- | --- |
| Within ambulance – rescue analgesia medication | Entonox, paracetamol, ibuprofen, other painkiller | Ambulance service data form | BNF 2021/22(45)  NHSSC 2021/22(46). |
| Within ambulance – side-effects medication | Midazolam or naloxone | Ambulance service data form | BNF 2021/22(45)  NHSSC 2021/22(46). |
| Inpatient care – index admission | Length of stay and number of days at each level of care | Hospital data collection form | NHS Reference Costs(44). |
| Index admission | CT scans | Hospital data collection form | NHS Reference Costs(44). |
| Inpatient care – follow up | Specified within CRFs | CRFs at 3m and 6m | NHS Reference Costs(44) and PSSRU(43). |
| Outpatient care | Specified within CRFs | CRFs at 3m and 6m | NHS Reference Costs(44) and PSSRU(43). |
| Community care | Specified within CRFs | CRFs at 3m and 6m | NHS Reference Costs(44) and PSSRU(43). |
| Medication | Specified within CRFs | CRFs at 3m and 6m | NHSBSA(47) |
| Personal social services | Specified within CRFs | CRFs at 3m and 6m | PSSRU(43). |

**Table S3: Unit Costs (£, UK) of resource inputs**

| Resource item | Unit cost | Unit | Source | Note |
| --- | --- | --- | --- | --- |
| **Emergency Response** |  |  |  |  |
| Intervention components |  |  |  |  |
| Morphine sulphate 10mg in 1ml ampoule | £0.63 | per ampoule | BNF 2021/22 |  |
| Ketamine hydrochloride 200mg in 20ml vial | £0.51 | per ampoule | BNF 2021/22 |  |
| Sodium chloride 0.9% 10ml ampoule | £1.39 | per pre-drawn syringe | BNF 2021/22 |  |
| 20g green intravenous cannula | £0.54 | per cannula | NHSSC 2021/22 |  |
| IV cannula dressing pack | £0.44 | per dressing pack | NHSSC 2021/22 |  |
| 10ml leur lock syringe | £0.12 | per syringe | NHSSC 2021/22 |  |
| Filter straw needle | £0.35 | per filter straw | NHSSC 2021/22 |  |
| Emergency ambulance | £3.90 | Per minute | National Schedule of NHS Costs 2021/22 | Cost per minute calculated by dividing cost of £390.08 by the average job cycle time (100mins) from the Operational productivity and performance in English NHS Ambulance Trusts publication. |
| Rescue Analgesia |  |  |  |  |
| Entonox | £5.21 | (EA size cylinder) | NHSSC 2021/22 | Includes the cost of a disposable filter |
| Paracetamol | £1.93 | 1g in 100ml infusion | BNF 2021/22 & NHSSC 2021/22 | Includes the cost of consumables |
| Ibuprofen | £0.07 | 2x200mg tablets | BNF 2021/22 |  |
| Side effects medication |  |  |  |  |
| Naloxone hydrochloride 400mcg in 1 ml ampoule | £5.31 | per ampoule | BNF 2021/22 & NHSSC 2021/22 | Includes the cost of consumables |
| Midazolam 2mg in 2ml ampoule | £2.06 | per ampoule | BNF 2021/22 & NHSSC 2021/22 | Includes the cost of consumables |
| **Emergency Department** |  |  |  |  |
| Attendance, not admitted | £183.91 | Per visit | National Schedule of NHS Costs 2021/22 | Weighted average non-admitted  (excluding dead on arrival) |
| Attendance, admitted | £265.65 | Per visit | National Schedule of NHS Costs 2021/22 | Weighted average admitted  (excluding dead on arrival) |
| **Critical Care** |  |  |  |  |
| XC01Z - XC07Z (Non-specific, general adult critical care) | £2,272.18 | Bed Day | National Schedule of NHS Costs 2021/22 | Weighted average cost of 0 organs to 6 or more organs supported. |
| **Inpatient admission** |  |  |  |  |
| Non-elective inpatient stays (long stays) | £4,974.00 | Per episode | Unit Costs of Health and Social Care 2021/22 |  |
| Non-elective inpatient stays (short stays) | £985.00 | Per episode | Unit Costs of Health and Social Care 2021/22 |  |
| **Outpatient visits** |  |  |  |  |
| Pain clinic | £204.36 | Per visit | National Schedule of NHS Costs 2021/22 |  |
| Outpatient clinic (other) | £123.89 | Per visit | National Schedule of NHS Costs 2021/22 |  |
| MRI scan | £45.77 | Per visit | National Schedule of NHS Costs 2021/22 |  |
| CT scan | £45.77 | Per visit | National Schedule of NHS Costs 2021/22 |  |
| X-ray | £45.77 | Per visit | National Schedule of NHS Costs 2021/22 |  |
| Ultrasound | £45.77 | Per visit | National Schedule of NHS Costs 2021/22 |  |
| A&E | £143.74 | Per visit | National Schedule of NHS Costs 2021/22 |  |
| Physiotherapy Service | £100.46 | Per visit | National Schedule of NHS Costs 2021/22 |  |
| General Surgery Service | £160.62 | Per visit | National Schedule of NHS Costs 2021/22 |  |
| Ear Nose and Throat Service | £155.17 | Per visit | National Schedule of NHS Costs 2021/22 |  |
| Orthopaedic Service | £157.16 | Per visit | National Schedule of NHS Costs 2021/22 |  |
| Gastroenterology Service | £148.93 | Per visit | National Schedule of NHS Costs 2021/22 |  |
| Orthopaedic Service | £157.16 | Per visit | National Schedule of NHS Costs 2021/22 |  |
| Neurology Service | £213.50 | Per visit | National Schedule of NHS Costs 2021/22 |  |
| Clinical Oncology Service | £160.43 | Per visit | National Schedule of NHS Costs 2021/22 |  |
| Orthotics Service | £171.70 | Per visit | National Schedule of NHS Costs 2021/22 |  |
| Cardiology Service | £169.39 | Per visit | National Schedule of NHS Costs 2021/22 |  |
| Plastic Surgery Service | £144.00 | Per visit | National Schedule of NHS Costs 2021/22 |  |
| Renal Medicine Service | £196.88 | Per visit | National Schedule of NHS Costs 2021/22 |  |
| Rheumatology Service | £165.17 | Per visit | National Schedule of NHS Costs 2021/22 |  |
| Podiatry Service | £93.368 | Per visit | National Schedule of NHS Costs 2021/22 |  |
| Ophthalmology Service | £141.97 | Per visit | National Schedule of NHS Costs 2021/22 |  |
| Urology Service | £137.73 | Per visit | National Schedule of NHS Costs 2021/22 |  |
| Anticoagulant service | £68.260 | Per visit | National Schedule of NHS Costs 2021/22 |  |
| **Community and social care** |  |  |  |  |
| GP surgery visits | £42 |  | Unit Costs of Health and Social Care 2021/22 |  |
| GP home visits | £100 |  | Unit Costs of Health and Social Care 2021/22 |  |
| GP telephone contacts | £8.80 |  | Unit Costs of Health and Social Care 2021/22 |  |
| GP video/online contacts | £14.43 |  | Unit Costs of Health and Social Care 2021/22 |  |
| District nurse contacts | £14.25 |  | Unit Costs of Health and Social Care 2021/22 | assumed to be 15 min consultation (Band 6 nurse) |
| Social worker contacts | £50 |  | Unit Costs of Health and Social Care 2021/22 |  |
| Physiotherapy contacts | £72 |  | Unit Costs of Health and Social Care 2021/22 | assumed 30 min appointment time |
| Occupational therapy contacts | £59 |  | Unit Costs of Health and Social Care 2021/22 | assumed 30 min appointment time |
| Counsellor | £58 |  | Unit Costs of Health and Social Care 2021/22 |  |
| Psychologist | £58 |  | Unit Costs of Health and Social Care 2021/22 |  |
| Home help/carer | £23 |  | Unit Costs of Health and Social Care 2021/22 |  |

**Table S4: Baseline demographic characteristics of participants by completeness of health economic data**

|  |  | Imputed (n=227) | Complete (n=189) |
| --- | --- | --- | --- |
| Age | Mean (SD) | 63.13 (23.49) | 63.62 (19.00) |
| Gender | Male n (%) | 111 (48.9%) | 85 (45.0%) |
|  | Female n (%) | 116 (51.1%) | 104 (55.0%) |
| Ethnicity | White | 122 (53.7%) | 76 (40.2%) |
|  | Black | - | - |
|  | Mixed | - | - |
|  | Any other ethnic group | 1 (0%) | 1 (1%) |
|  | Asian | - | 1 (1%) |
|  | Ethnicity not given | 104 (45.8%) | 110 (58.2%) |
|  | Missing | - | 1 (1%) |
| Weight | Mean (SD) | 74.42 (15.93) | 79.40 (20.96) |
| Initial pain score | Mean (SD) | 8.91 (1.15) | 8.76 (1.26) |

**Table S5: Health-related quality of life, resource use and cost outcomes for observed data.**

|  | **Ketamine** | | **Morphine** | | **Difference** | |
| --- | --- | --- | --- | --- | --- | --- |
|  | **(N), mean** | **(SD)** | **(N), mean** | **(SD)** | **mean** | **(p value)** |
| **Health-related quality of life** |  |  |  |  |  |  |
| EQ-5D Baseline | (206), 0.7809 | (0.07) | (210), 0.7818 | (0.08) | -0.0009 | 0.9059 |
| EQ-5D 3 months | (114), 0.5395 | (0.31) | (111), 0.5052 | (0.29) | 0.0343 | 0.3874 |
| EQ-5D 6 months | (107), 0.5925 | (0.30) | (115), 0.5324 | (0.34) | 0.0601 | 0.1608 |
| QALYs | (98), 0.3092 | (0.10) | (98), 0.2931 | (0.10) | 0.0161 | 0.2733 |
| **Resource use** (all visits) |  |  |  |  |  |  |
| Inpatient nights | (103), 0.5631 | (3.58) | (104), 1.4615 | (8.36) | -0.8984 | 0.3160 |
| Outpatient |  |  |  |  |  |  |
| Pain clinic | (2), 1.50 | (0.71) | (1), 3 | (0) | NA | NA |
| Outpatient clinic (other) | (68), 3.44 | (3.39) | (59), 2.51 | (2.06) | 0.9327 | 0.0685 |
| MRI scan | (9), 1.33 | (0.50) | (12), 1.17 | (0.39) | 0.1667 | 0.4003 |
| CT scan | (9), 1.22 | (0.44) | (11), 1.18 | (0.40) | 0.0404 | 0.8334 |
| X-ray | (46), 2.33 | (1.76) | (40), 2.25 | (1.61) | 0.0761 | 0.8361 |
| Ultrasound | (4), 2.25 | (1.89) | (7), 1 | (0) | 1.2500 | 0.1013 |
| A&E | (6), 1.67 | (1.21) | (10), 1.1 | (0.32) | 0.5667 | 0.1744 |
| Other visits | (71), 3.67 | (3.28) | (59), 4.05 | (4.44) | -0.3748 | 0.5816 |
| Community |  |  |  |  |  |  |
| GP surgery visits | (31), 2.45 | (1.79) | (18), 1.50 | (0.79) | 0.9516 | 0.0379 |
| GP home visits | (7), 4.57 | (6.45) | (8), 2 | (1.93) | 2.5714 | 0.3003 |
| GP telephone contacts | (28), 2.39 | (1.75) | (41), 1.78 | (1.04) | 0.6124 | 0.0727 |
| GP video/online contacts | (5), 8.80 | (9.26) | (1), 1 | (0) | 7.8000 | NA |
| District nurse contacts | (22), 7.91 | (7.58) | (12), 4.25 | (4.00) | 3.6591 | 0.1309 |
| Social worker contacts | (4), 2.00 | (2.00) | (6), 1.17 | (0.41) | 0.8333 | 0.3379 |
| Physiotherapy contacts | (81), 4.64 | (4.32) | (53), 4.98 | (5.46) | -0.3392 | 0.6901 |
| Occupational therapy contacts | (10), 2.30 | (1.34) | (19), 3.68 | (6.60) | -1.3842 | 0.5207 |
| Counsellor | (1), 2 | (0) | (0), 0 | (0) |  | NA |
| Psychologist | (5), 3.20 | (2.49) | (0), 0 | (0) |  | NA |
| Home help/carer | (9), 36.56 | (44.19) | (20), 70.40 | (89.85) | -33.8444 | 0.2960 |
| Other community contacts | (13), 7 | (7.69) | (12), 5.25 | (6.59) | 1.7500 | 0.5490 |
| **Costs (£)** |  |  |  |  |  |  |
| Treatment | (206), 21.76 | (10.17) | (210), 23.89 | (11.92) | -2.1347 | 0.0499 |
| Ambulance | (206), 330.72 | (94.65) | (210), 343.34 | (110.80) | -12.6221 | 0.2120 |
| Hospital admission | (206), 3547.65 | (3169.76) | (210), 3141.72 | (2500.16) | 405.9300 | 0.1483 |
| Hospital readmission | (103), 151.95 | (741.75) | (104), 511.46 | (1884.43) | -359.5081 | 0.0727 |
| Outpatient | (102), 582.23 | (889.60) | (103), 417.65 | (600.26) | 164.5790 | 0.1226 |
| NHS Medication | (103), 13.49 | (22.01) | (100), 11.63 | (26.18) | 1.8621 | 0.5845 |
| Community & social | (101), 429.63 | (637.34) | (103), 549.37 | (1273.55) | -119.7426 | 0.3958 |
| NHS equipment | (101), 103.65 | (137.78) | (102), 74.57 | (100.86) | 29.0779 | 0.0882 |
| TOTAL NHS&PSS | (99), 5191.42 | (3154.51) | (97), 5143.98 | (3897.15) | 47.4341 | 0.9255 |
| Wider/societal | (101), 1192.29 | (4570.41) | (101), 1110.32 | (3691.16) | 81.9703 | 0.8886 |
| Non-NHS Medication | (103), 3.65 | (8.00) | (100), 2.92 | (5.00) | 0.7275 | 0.4369 |
| Non-NHS equipment | (101), 45.20 | (247.35) | (102), 51.39 | (380.21) | -6.1961 | 0.8906 |
| Total Non-NHS Cost | (100), 1252.59 | (4601.64) | (99), 1142.35 | (3728.68) | 110.2339 | 0.8528 |
| **Total NHS and Non-NHS** | (98), 6441.79 | (5627.60) | (97), 6309.89 | (6010.68) | 131.9021 | 0.8745 |

**Table S6: Cost-effectiveness results, mean costs, and effects**

|  | COSTS, mean (SE) | | QALYS, mean (SE) | |
| --- | --- | --- | --- | --- |
|  | Ketamine | Morphine | Ketamine | Morphine |
| Base case | £5207 (289.20) | £5324 (324.09) | 0.314 (0.01) | 0.289 (0.01) |
| Sensitivity analyses |  |  |  |  |
| Inclusion of societal costs | £6266 (417.08) | £6373 (467.83) | 0.314 (0.01) | 0.289 (0.01) |
| Complete case | £5317 (350.87) | £5084 (352.75) | 0.309 (0.01) | 0.294 (0.01) |
| Baseline utility assumptions changes | £5207 (289.20) | £5324 (324.09) | 0.216 (0.01) | 0.191 (0.01) |
| Subgroup analyses |  |  |  |  |
| Age <60 | £4654 (449.87) | £3862 (512.21) | 0.311 (0.02) | 0.291 (0.02) |
| Age ≥60 | £5566 (381.55) | £6288 (425.83) | 0.316 (0.01) | 0.287 (0.01) |
| Female | £5180 (405.72) | £5192 (447.85) | 0.292 (0.01) | 0.283 (0.01) |
| Male | £5236 (431.56) | £5471 (486.52) | 0.339 (0.01) | 0.295 (0.01) |
| Analgesia, no | £4975 (381.46) | £5450 (408.62) | 0.319 (0.01) | 0.295 (0.01) |
| Analgesia, yes | £5526 (452.55) | £5147 (501.85) | 0.307 (0.01) | 0.280 (0.01) |
|  | COSTS, mean (SE) | | Effects, mean (SE) | |
| Scenario analysis |  |  |  |  |
| Cost per unit change in SPID score | £3922 (193.70) | £3485 (189.94) | 3.436 (0.20) | 3.338 (0.19) |
